# Public Exposure to Live Animals, Behavioural Change, and Support in Containment Measures in response to COVID-19 Outbreak: a population-based cross sectional survey in China

**DOI:** 10.1101/2020.02.21.20026146

**Authors:** Zhiyuan Hou, Leesa Lin, Lu Liang, Fanxing Du, Mengcen Qian, Yuxia Liang, Juanjuan Zhang, Hongjie Yu

## Abstract

**Background:** In response to the COVID-19 outbreak, we aimed to investigate behavioural change on exposure to live animals before and during the outbreak, and public support and confidence for governmental containment measures.

**Methods:** A population-based cross-sectional telephone survey via random dialing was conducted in Wuhan (the epicentre) and Shanghai (an affected city with imported cases) between 1 and 10 February, 2020. 510 residents in Wuhan and 501 residents in Shanghai were randomly sampled. Differences of outcome measures were compared before and during the outbreak, and between two cities.

**Findings:** Proportion of respondents visiting wet markets at usual was 23.3% (119/510) in Wuhan and 20.4% (102/501) in Shanghai. During the outbreak, it decreased to 3.1% (16) in Wuhan (p<0·001), and 4.4% (22) in Shanghai (p<0·001). Proportion of those consuming wild animal products declined from 10.2% (52) to 0.6% (3) in Wuhan (p<0·001), and from 5.2% (26) to 0.8% (4) in Shanghai (p<0·001). 79.0% (403) of respondents in Wuhan and 66.9% (335) of respondents in Shanghai supported permanent closure of wet markets (P<0.001). 95% and 92% of respondents supported banning wild animal trade and quarantining Wuhan, and 75% were confident towards containment measures. Females and the more educated were more supportive for the above containment measures.

**Interpretation:** The public responded quickly to the outbreak, and reduced exposure to live animals, especially in Wuhan. With high public support in containment measures, better regulation of wet markets and healthy diets should be promoted.

**Funding:** National Science Fund for Distinguished Young Scholars, H2020 MOOD project.

**Research in context:** *Evidence before this study:* On February 19, 2020, we searched PubMed for papers published after January 1, 2020, containing the following terms: “2019 nCoV” or “COVID-19”. We identified 179 studies, most of which are research on clinical and epidemiological characteristics of COVID-19. To date there is no primary research to quantify public behavioural response and support in containment measures in response to the outbreak. Only four commentaries mentioned the influence of the outbreak on mental health. One commentary introduced the habit of consuming wild animal products in China. Another commentary briefly introduced isolation, quarantine, social distancing and community containment as public health measures in the outbreak. The Chinese government has introduced a series of strict containment measures, and societal acceptability of these measure is important for effective and sustained response. Evidence is urgently needed to help policy makers understand public response to the outbreak and support for the containment measures, but no evidence available to date.

*Added value of this study:* We conducted a population-based cross-sectional telephone survey via random digital dialing in Wuhan (the epicentre) and Shanghai (an affected city with imported cases) between 1 and 10 February, 2020. To date, this is the only few analyses on behavioural response to the outbreak and societal acceptability of governmental containment measures, which has been listed as the current priority of China CDC. We provide an assessment of behavioural change on exposure to live animals during the outbreak, by comparison before and during the outbreak, and between two cities with diverse exposure intensities to COVID-19. We also provide evidence on public support in governmental containment measures, including strict regulation on wet markets to reduce animal-to-human transmission and city quarantine to reduce human transmission.

*Implications of all the available evidence:* We found that wild animal consumption was more prevalent in Wuhan (10.2%) than in Shanghai (5.2%). The public responded quickly to the outbreak, and significantly reduced exposure to live animals and stopped wild animal consumption, especially in Wuhan. They were very supportive of governmental containment measures. With high public support, wet markets should be better regulated, and healthy diets, including changing the traditional habit of eating wild animal products, should be promoted. This can inform policy makers in China and other countries to implement and adjust containment strategies in response to the outbreak in the future.

## Introduction

Early in December 2019, several pneumonia cases caused by the novel coronavirus (now known as COVID-19) were first reported in Wuhan city, China.^1-3^ The majority of the earliest cases were epidemiologically traced to the local Huanan (Southern China) Seafood Wholesale Market, where some wild animals were sold.^1-4^ On Jan 26, novel coronavirus was detected in environmental samples from this market by China CDC, suggesting that the virus possibly originated in wild animals sold in the market.^3-4^ This pathogenic virus has been identified as a new strain of coronavirus, which is in the same family as severe acute respiratory syndrome coronavirus (SARS-CoV) and Middle East respiratory syndrome coronavirus (MERS-CoV).^4-5^ The COVID-19 outbreak was declared by World Health Organization as a Public Health Emergency of International Concern (PHEIC) on Jan 30, 2020.^6^ As of Feb 16, 2020, a total of 70,548 cases have been reported in mainland China, with 1,770 resulting deaths.^7^ The outbreak has now spread beyond China to twenty-five other countries.^8^

The SARS-CoV and MERS-CoV were proven to have originated from wild animals.^9-14^ The COVID-19 is also likely to have a zoonotic origin. The practice of consuming wild animal products in China dates back to prehistoric times and persists into today.^9^ Trade and consumption of wild animals were not well regulated even after SARS. Until the amendment of Wild Animal Protection Act in 2016, the prohibition was firstly added, but only for a short list of the nationally protected animals. Previous studies explored population exposure to live poultry in China,^15-17^ but information on exposure to wild animals is limited.

In the efforts to contain the spread of the virus, the Chinese government introduced a series of containment measures. ^18^ Specifically, to lower the risk of zoonotic virus transmission, China has issued strict regulations on wet markets where live and/or wild animals are traded, including closing live poultry markets (LPMs) and bans on wild animal transactions. Furthermore, Wuhan city has been quarantined to stop human-to-human transmission: starting on Jan 23, 2020, public transportation in Wuhan was closed, and all flights and trains leaving from Wuhan were cancelled.

The COVID-19 outbreak and subsequent containment measures to lower the zoonotic virus transmission offer a unique opportunity to reassess the exposure to live animals, demand for wild animal consumption, and public readiness to further regulate wet markets. In this study, we aim to investigate behaviour patterns of population interaction to live animals, with a particular focus on differences before and during the outbreak, between Wuhan and Shanghai representing diverse exposure intensities to COVID-19. We also seek to understand levels of public support for containment measures and public confidence in their effectiveness to quell the outbreak. This study will help to clarify the public’s response to the outbreak and has important policy implications regarding the social acceptability of infection containment measures and their application in different contexts.

## Methods

### Study design

A population-based cross-sectional telephone survey was conducted in Wuhan and Shanghai between Feb 1 and 10, 2020, one month since the outbreak. These two cities were selected to represent diverse exposure intensities to COVID-19. Wuhan is the site of origin of the COVID-19 outbreak. Shanghai was selected due to its status as a city that has been significantly affected by imported COVID-19 cases from Wuhan. The first imported case was confirmed in Shanghai on Jan 20. During the period of our survey, the number of confirmed cases rose rapidly in Wuhan, from 4,109 to 18,454, but maintained low epidemic rates in Shanghai, with reported cases rising from 182 to 303. Figure 1 illustrates the timeline of the outbreak progression and containment measures. This provides an opportunity to assess and compare public behavioural response and support for containment measures during the outbreak between two cities with different exposure intensities.

**Figure 1.**
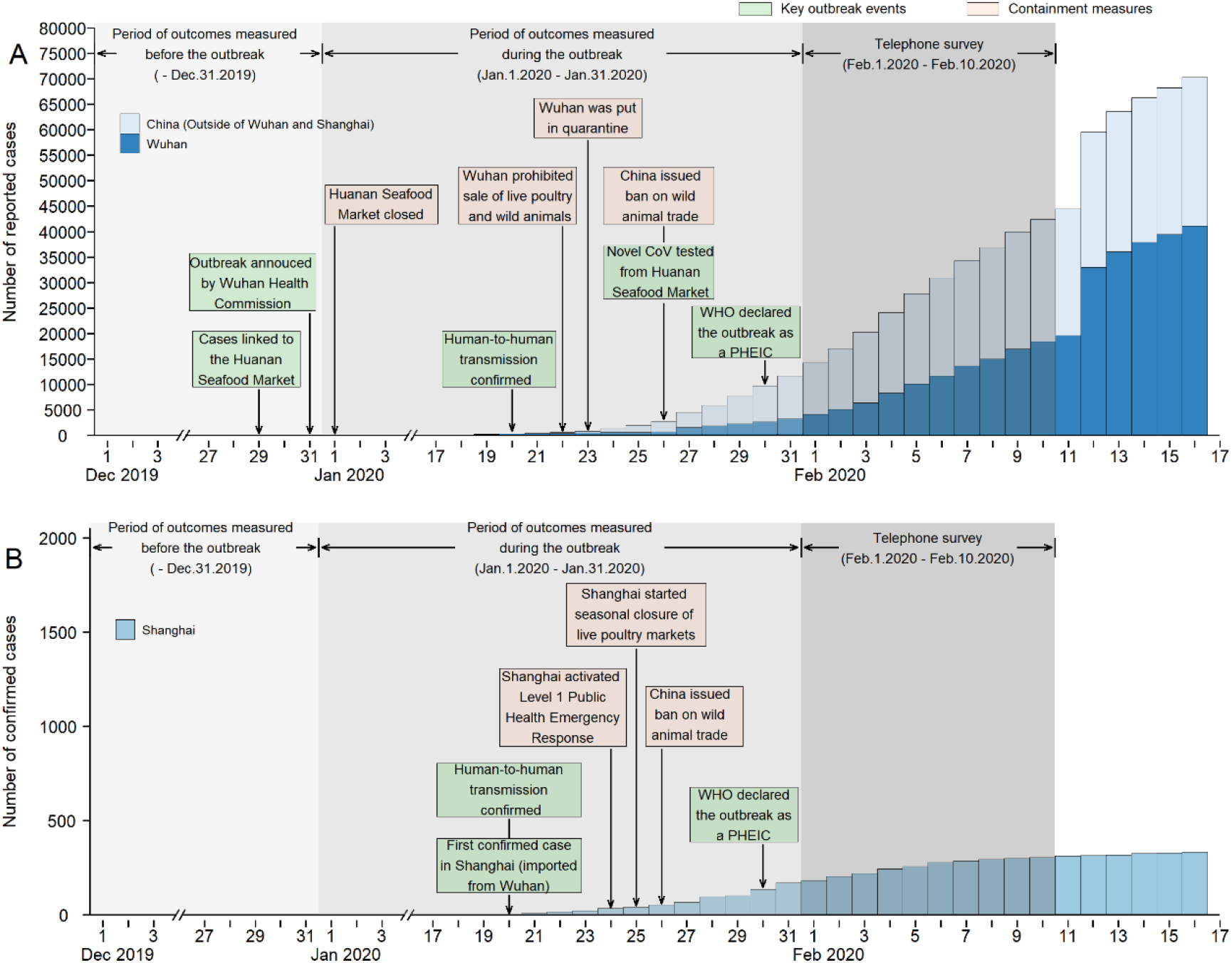
The timeline of the COVID-19 outbreak and containment measure. Notes: **Panel A** for Wuhan and other parts of China and **Panel B** for Shanghai, and scales are different in two panels. The vertical axis for Panel A is number of reported cases and that for Panel B is number of confirmed cases, because Wuhan changed diagnostic criteria on Feb 12, 2020, and since then confirmed cases are based on clinical diagnosis instead of laboratory testing. Outcomes were measured in two periods (before and during the outbreak). Key outbreak events and containment measures are listed in different colours. WHO stands for World Health Organization; PHEIC stands for public health emergency of international concern.

The sample size (n=500 for each city) is sufficient to estimate the change of outcomes (p=0.5) with a margin of error of 0.05 and a 95% confidence interval, according to previous studies.^15-16^ Our target population was persons aged 18 or over who had been living in the selected city for at least 6 months in the year prior to the survey and were current residents at the time of the survey. In each city, the telephone survey was conducted through a computer-assisted interviewing system, which enabled the random generation of mobile phone numbers and systematic data collection. We attempted to contact each generated number three times a day at different hours; if no contact was established after the third call, this number was classified as invalid or unreachable. Random dials with proportional quota sampling were used to ensure that respondents were demographically representative of the general population in each city, with quotas based on sex and age.^19^

### Measures

The outcomes of interest we studied were exposure to live animals before and during the COVID-19 outbreak, public support for containment measures that aim to reduce population exposure to live animals and human transmission, and confidence in these measures to quell the outbreak. Exposure to live animals includes the following behaviours: visiting the live poultry or wild animal markets, buying live poultry or wild animals (i.e., frequency of purchases, practice of picking up live animals before purchasing, location where the purchased animals were slaughtered), eating wild animal products, raising pets, and touching stray animals. To measure behavioural change during the outbreak, each respondent was asked to report exposing behaviours both (1) prior to the outbreak: before Dec 31, 2019 when COVID-19 cases were first released by Wuhan Health Commission and (2) since the outbreak, between Jan 1 to Feb 10, 2020. For live animal consumption habits, where behaviour change cannot be measured in a short period of time, we assessed people’s intention to: 1) buy live poultry or wild animals, and eat wild animal products (continue to buy/eat as usual, buy/eat less, not buy/eat any more) and 2) continue raising pets and touching stray animals (continue to raise/touch as usual, temporarily not during the outbreak, try or resolutely not even after the outbreak).

We measured public support for two types of governmental containment measures in response to the outbreak; one is regulation on wet markets to reduce population exposure to live animals, and the other is quarantining measures meant to reduce human transmission, such as the closure of public transportation in Wuhan and the cancellation of flights and trains leaving from Wuhan. Each respondent was asked to rate the extent, on a five-point Likert scale, to which they supported 1) the permanent closure of LPMs, 2) a ban on the wild animal trade, and 3) the quarantining of Wuhan; respondents were also asked to report the extent of their confidence in the effectiveness of these measures to quell the outbreak.

Characteristics of respondents, including gender, age, marital status, education, income, and employment status, were collected in our survey. We also collected the history of fever or cough symptoms for respondents and their relatives or friends in the two weeks prior to the survey, and whether there were confirmed or suspected COVID-19 cases in the neighbourhood in which they live. Each respondent was also asked at what point they started paying attention to this outbreak.

The survey questionnaire was adjusted according to an instrument used during the outbreaks of SARS in 2003 ^20-21^ and influenza A(H7N9) in 2013.^15^ The survey instrument was pretested for face and content validity, length, and understandability by the public before our survey was administered.

### Statistical Analysis

Descriptive and univariate analysis were used to investigate changes in exposure to live animals during the outbreak, public support for containment measures, and confidence in containment measures to quell the outbreak, between Wuhan (the epicentre) and Shanghai (an affected city with imported cases). Chi-square tests or fisher’s exact tests (when applicable) were used to compare differences in these measures between the two cities, and before and during the outbreak. Multivariate logistic regression was used to estimate factors associated with public support for containment measures and confidence in intervention effectiveness. The proportion and Odds Ratio with 95% confidence intervals (CI) were reported. All statistical analyses were performed using Stata 14.0 (StataCorp LP, College Station, TX).

## Results

A total of 18,576 telephone numbers were randomly selected via digital dialing system in Wuhan and Shanghai. After calling, 6,836 respondents were eligible to participate, of whom 5,640 declined and 185 terminated before completing the interview. In total, 510 respondents in Wuhan and 501 respondents in Shanghai completed the interview, and the overall response rate was 14.8% (1,011/6,836) (Appendix Figure 1).

Respondents in Wuhan and Shanghai were generally similar in terms of sex, marital status and occupation, but in Shanghai respondents tended to be older and had higher education and income levels than in Wuhan (Table 1). Our sample was consistent with characteristics of the general population in the two cities, as Shanghai is more developed and has an older population than Wuhan. Expectedly, respondents in Wuhan were much more intensely exposed to the COVID-19 outbreak and paid attention to the outbreak earlier than those in Shanghai.

**Table 1.** Characteristics of participants and COVID-19 exposure at Wuhan and Shanghai, China.

|  | Wuhan, (n=510) | Shanghai, (n=501) | P value |
| --- | --- | --- | --- |
| **Sociodemographic characteristics** |  |  |  |
| Sex |  |  | 0.635 |
| Female | 254(49.8) | 257(51.3) |  |
| Male | 256(50.2) | 244(48.7) |  |
| Age group, years |  |  | 0.049 |
| 18-24 | 89(17.5) | 75(15.0) |  |
| 25-39 | 161(31.6) | 176(35.1) |  |
| 40-59 | 197(38.6) | 165(32.9) |  |
| 60+ | 63(12.4) | 85(17.0) |  |
| Marital status |  |  | 0.051 |
| Single | 122(23.9) | 147(29.3) |  |
| Married | 388(76.1) | 354(70.7) |  |
| Education |  |  | <0.001 |
| Middle school or below | 29(5.7) | 51(10.2) |  |
| High school | 110(21.6) | 75(15.0) |  |
| Three years college | 191(37.5) | 108(21.6) |  |
| Bachelor or above | 180(35.3) | 267(53.3) |  |
| Household income level |  |  | 0.009 |
| Low | 122(23.9) | 97(19.4) |  |
| Middle | 322(63.1) | 306(61.1) |  |
| High | 66(12.9) | 98(19.6) |  |
| Employment |  |  | 0.806 |
| Retired/unemployed | 140(27.4) | 141(28.1) |  |
| Employed | 370(72.6) | 360(71.9) |  |
| **The COVID-19 exposure** |  |  |  |
| Experienced symptoms* | 63(12.4) | 51(10.2) | 0.275 |
| Relatives or friends with symptoms* | 64(12.6) | 36(7.2) | 0.004 |
| Suspected cases in neighbourhood |  |  | <0.001 |
| Yes | 175(34.3) | 32(6.4) |  |
| No | 260(51.0) | 413(82.4) |  |
| Unclear | 75(14.7) | 56(11.2) |  |
| Day of paying attention to the outbreak |  |  | <0.001 |
| Before Dec 31, 2019 | 94(18.4) | 44(8.8) |  |
| Jan 1-12, 2020 | 127(24.9) | 72(14.4) |  |
| Jan 13-19, 2020 | 70(13.7) | 64(12.8) |  |
| Jan 20-22, 2020 | 111(21.8) | 161(32.1) |  |
| After Jan 23, 2020 | 108(21.2) | 160(31.9) |  |

All data are n (%). P value are from Chi square test or fisher's exact tests (when applicable).
\*Fever or cough symptoms in the past 2 weeks.

We first investigated patterns of usual exposure to live animals before the outbreak (Table 2 Panel A). Respondents reported visiting LPMs or buying live animals at similar rates in the two cities before the outbreak, although wet markets in Wuhan were significantly less accessible than in Shanghai. In both cities, approximately one in five respondents reported having visited LPMs in the year prior to the outbreak (119, 23.3% in Wuhan vs 102, 20.4% in Shanghai, p=0.210), and about 11% reported buying live poultry or wild animals from wet markets. Among those who bought live animals, about half reported that they picked up animals for examination before buying (27 of 55, 49.1% in Wuhan vs 32 of 55, 58.2% in Shanghai, p=0.339); more than 90% of respondents always arranged for slaughter of purchased animals in wet markets, with no noticeable differences between the two cities. However, residents in Wuhan reported wild animal consumption at a higher rate than those in Shanghai; 47 (9.2%) of 510 respondents in Wuhan and 23 (4.6%) of 501 respondents in Shanghai reported eating wild animal products (p=0.004), and 19 (3.7%) respondents in Wuhan and 7 (1.4%) respondents in Shanghai had visited wild animal markets in the year prior to the outbreak. Fewer respondents in Wuhan raised pets (69, 13.5% vs 95, 19.0%; p=0.019) or touched stray animals (43, 8.4% vs 58, 11.6%; p=0.095) than in Shanghai.

**Table 2:**
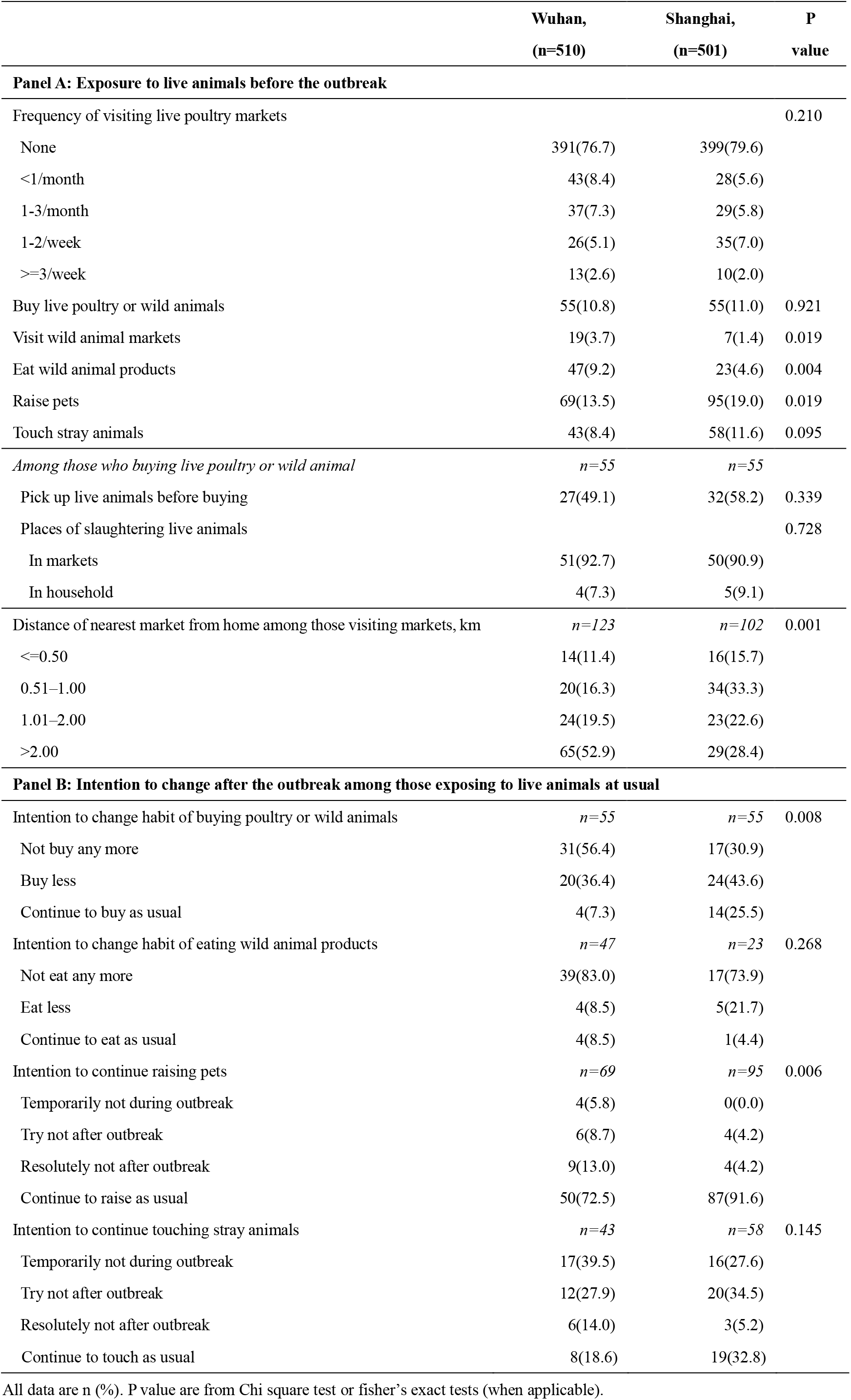
Exposure to live animals before and intention to change after the COVID-19 outbreak, China.

|  | Wuhan,<br>(n=510) | Shanghai,<br>(n=501) | P<br>value |
| --- | --- | --- | --- |
| **Panel A: Exposure to live animals before the outbreak** |  |  |  |
| Frequency of visiting live poultry markets |  |  | 0.210 |
| None | 391(76.7) | 399(79.6) |  |
| <1/month | 43(8.4) | 28(5.6) |  |
| 1-3/month | 37(7.3) | 29(5.8) |  |
| 1-2/week | 26(5.1) | 35(7.0) |  |
| >=3/week | 13(2.6) | 10(2.0) |  |
| Buy live poultry or wild animals | 55(10.8) | 55(11.0) | 0.921 |
| Visit wild animal markets | 19(3.7) | 7(1.4) | 0.019 |
| Eat wild animal products | 47(9.2) | 23(4.6) | 0.004 |
| Raise pets | 69(13.5) | 95(19.0) | 0.019 |
| Touch stray animals | 43(8.4) | 58(11.6) | 0.095 |
| *Among those who buying live poultry or wild animal* | *n=55* | *n=55* |  |
| Pick up live animals before buying | 27(49.1) | 32(58.2) | 0.339 |
| Places of slaughtering live animals |  |  | 0.728 |
| In markets | 51(92.7) | 50(90.9) |  |
| In household | 4(7.3) | 5(9.1) |  |
| Distance of nearest market from home among those visiting markets, km | *n=123* | *n=102* | 0.001 |
| <=0.50 | 14(11.4) | 16(15.7) |  |
| 0.51–1.00 | 20(16.3) | 34(33.3) |  |
| 1.01–2.00 | 24(19.5) | 23(22.6) |  |
| >2.00 | 65(52.9) | 29(28.4) |  |
| **Panel B: Intention to change after the outbreak among those exposing to live animals at usual** |  |  |  |
| Intention to change habit of buying poultry or wild animals | *n=55* | *n=55* | 0.008 |
| Not buy any more | 31(56.4) | 17(30.9) |  |
| Buy less | 20(36.4) | 24(43.6) |  |
| Continue to buy as usual | 4(7.3) | 14(25.5) |  |
| Intention to change habit of eating wild animal products | *n=47* | *n=23* | 0.268 |
| Not eat any more | 39(83.0) | 17(73.9) |  |
| Eat less | 4(8.5) | 5(21.7) |  |
| Continue to eat as usual | 4(8.5) | 1(4.4) |  |
| Intention to continue raising pets | *n=69* | *n=95* | 0.006 |
| Temporarily not during outbreak | 4(5.8) | 0(0.0) |  |
| Try not after outbreak | 6(8.7) | 4(4.2) |  |
| Resolutely not after outbreak | 9(13.0) | 4(4.2) |  |
| Continue to raise as usual | 50(72.5) | 87(91.6) |  |
| Intention to continue touching stray animals | *n=43* | *n=58* | 0.145 |
| Temporarily not during outbreak | 17(39.5) | 16(27.6) |  |
| Try not after outbreak | 12(27.9) | 20(34.5) |  |
| Resolutely not after outbreak | 6(14.0) | 3(5.2) |  |

|  |  |  |
| --- | --- | --- |
| Continue to touch as usual | 8(18.6) | 19(32.8) |

All data are n (%). P value are from Chi square test or fisher's exact tests (when applicable).

To investigate effects of the outbreak, we further assessed behavioural changes in exposure to live animals before and during the outbreak (Figure 2), and intention to change after the outbreak (Table 2 Panel B). We found that respondents dramatically reduced exposure to live animals during the outbreak, with respondents in Wuhan changing their exposure much more significantly than in Shanghai. Specifically, the frequency of visiting LPMs decreased from 23.3% (n=119) to 3.1% (n=16) in Wuhan (p<0·001), and from 20.4% (n=102) to 4.4%(n=22) in Shanghai (p<0·001; Figure 2). The proportion of respondents buying live poultry or wild animals also decreased from around 11.0% to 2.6% in both cities (p<0·001). We also asked whether respondents intended to change their habit of buying live poultry or wild animals since the outbreak among those exposing to live animals at usual (Table 2 Panel B). In Wuhan and Shanghai, 56.4% (31/55) and 30.9% (17/55) of respondents claimed to have stopped buying live poultry or wild animals, respectively. 36.4% (20/55) of respondents in Wuhan and 43.6% (24/55) in Shanghai claimed they intended to buy less, whereas the remaining 7.3% (4/55) of respondents in Wuhan and 25.5% (14/55) in Shanghai stated they would continue to buy these products as usual.

**Figure 2:**
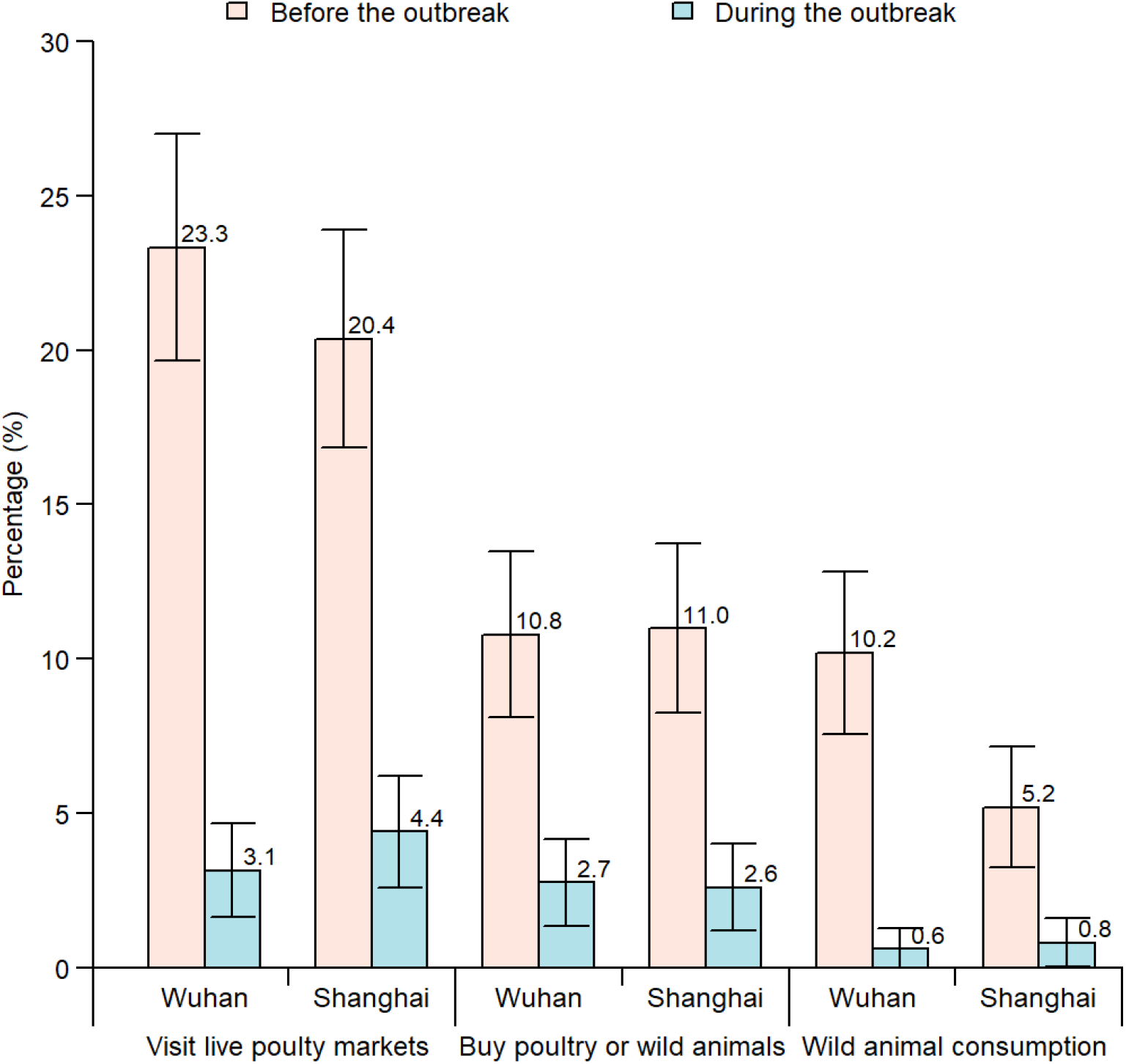
Proportion of respondents exposing to live animals before and after the COVID-19 outbreak, China. Notes: Proportions are shown, and error bars represent 95% CIs.

The proportion of respondents consuming wild animals declined significantly, 10.2%(n=52) to 0.6%(n=3) in Wuhan (p<0·001) and 5.2%(n=26) to 0.8%(n=4) in Shanghai (p<0·001) (Figure 2). Most respondents reported having stopped consuming wild animals since the outbreak. However, the behaviour of raising pets did not change much, with 72.5% (50/69) and 91.6% (87/95) of respondents claiming to continue raising pets as usual in Wuhan and Shanghai, respectively (Table 2 Panel B). 39.5% (17/43) and 27.6% (16/58) of respondents in Wuhan and Shanghai, respectively, claimed that they would temporarily not touch stray animals during the outbreak, and 41.9% (18/43) and 39.7% (23/58), respectively, would try not to or definitely not touch stray animals, even after the outbreak. The remaining 18.6% (8/43) and 32.8% (19/58) of respondents in Wuhan and Shanghai, respectively, would continue to touch stray animals as usual.

Figure 3 displays responses regarding public support for containment measures and confidence in these measures. 403 (79.0%) of 510 respondents in Wuhan and 335 (66.9%) of 501 respondents in Shanghai claimed to support the permanent closure of LPMs to quell the outbreak (P<0.001). In both cities, 95% and 92% of respondents supported banning the wild animal trade and the policy of quarantining Wuhan, respectively, and 75% of respondents were confident that governmental containment measures would quell the outbreak.

**Figure 3:**
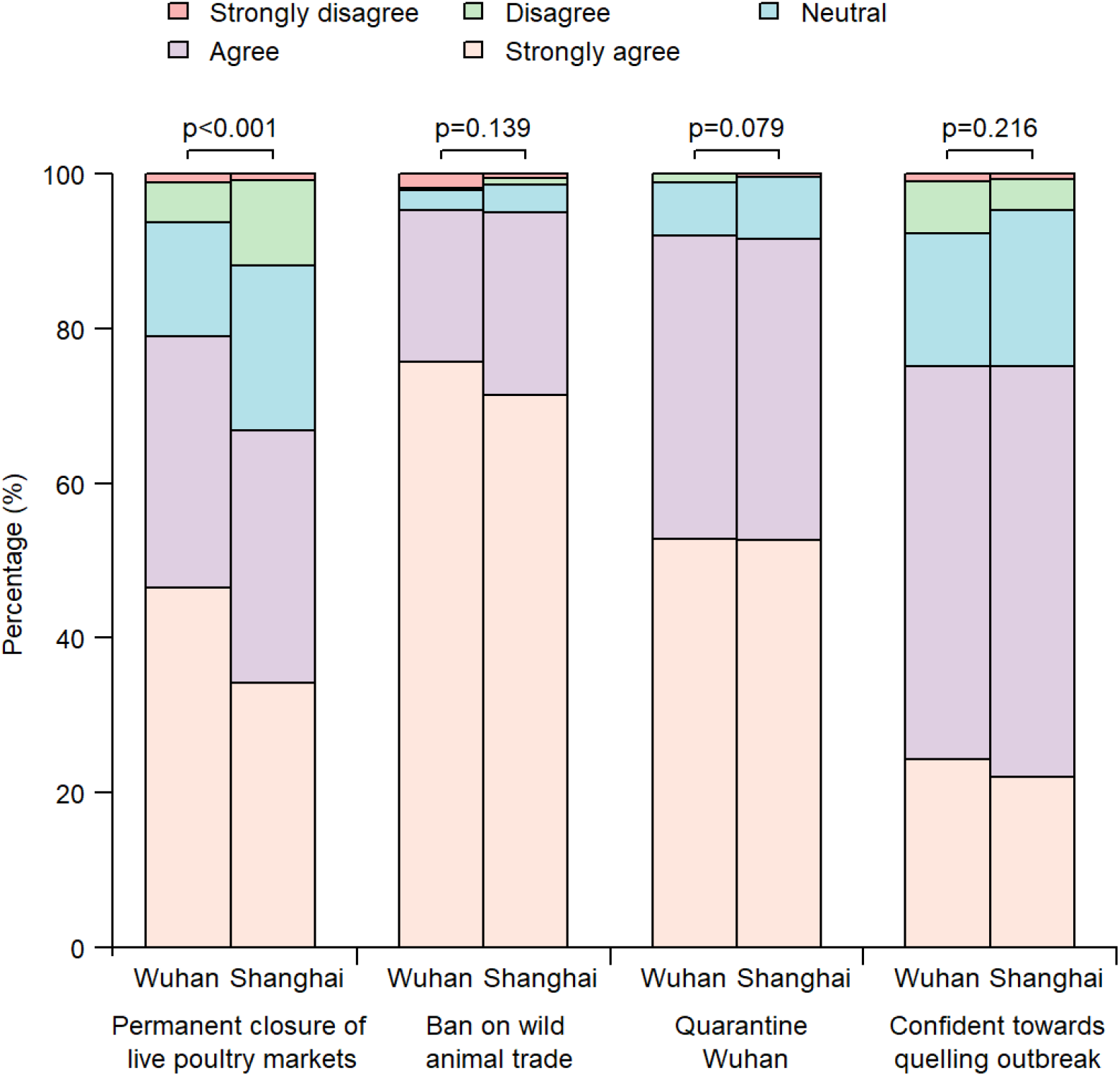
Public support and confidence for containment measures in response to the COVID-19 outbreak in China. Notes: p value is shown between Wuhan and Shanghai.

Table 3 presents socio-demographic factors associated with public support for the governmental containment measures implemented (i.e. closure of LPMs, banning the wild animal trade, and city quarantine) and confidence in the effectiveness of governmental response. There was evidence that females were more supportive towards all three containment measures and that there was a positive association between education level and the likelihood of supporting regulations on wet markets and trade of wild animals. Unsurprisingly, those who self-reported visiting a wet market and eating wild animal products prior to the outbreak are much less likely to support closure of the market or a ban on the trade as containment measures (adjusted OR=0.58, 95%CI [0.41-0.82] and adjusted OR=0.31, 95%CI [0.14-0.70], respectively). We did not find any difference in population profiles of those who support Wuhan quarantine or show confidence in the effectiveness of governmental response.

**Table 3:**
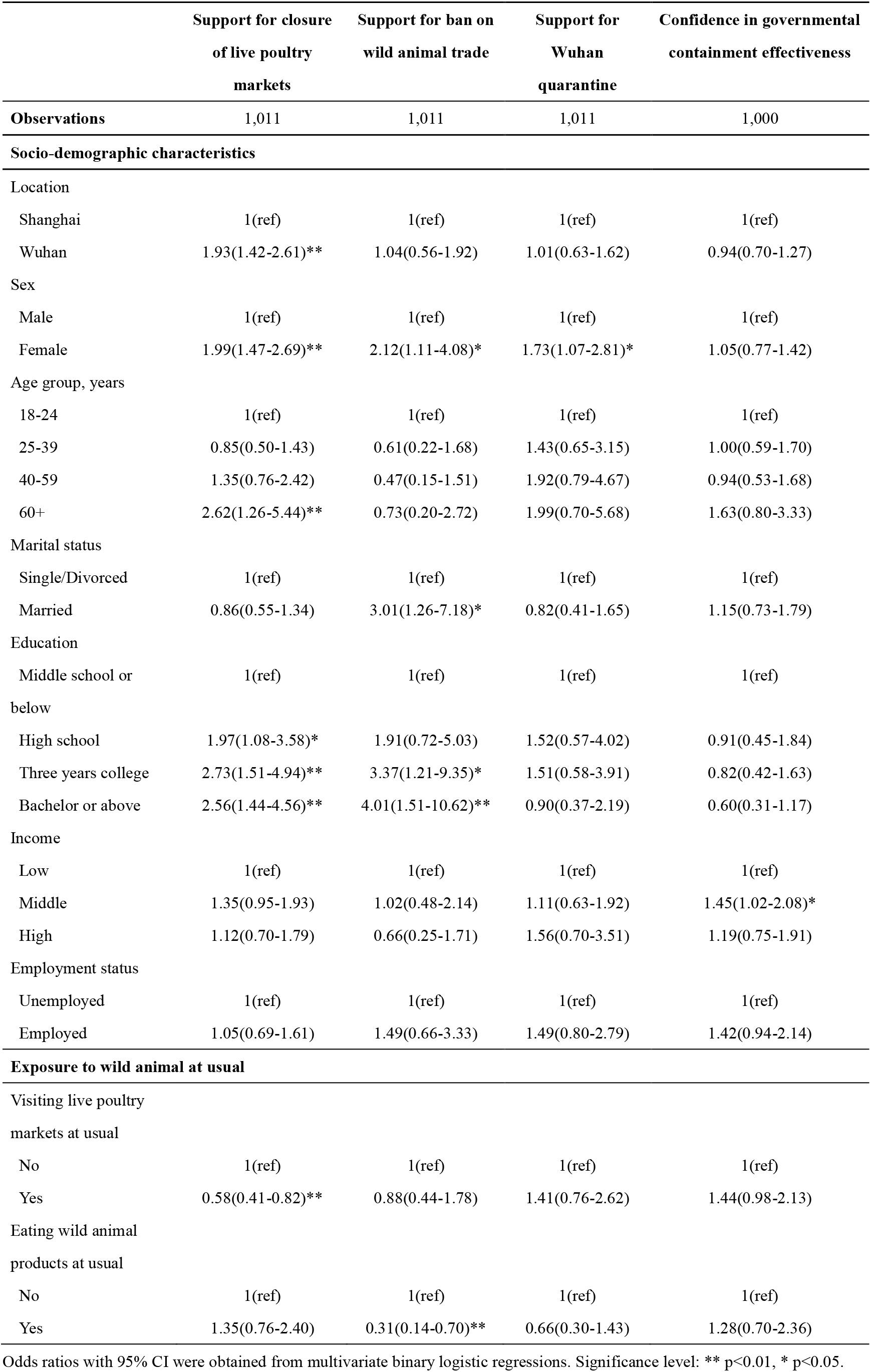
Factors associated with support and confidence for containment measures in response to the COVID-19 outbreak in China.

## Discussion

This study is the first to investigate behavioural changes of exposure to live animals during the COVID-19 outbreak, and public support for governmental containment measures in response to the outbreak. We found that population exposures to live animals were similar in Wuhan and Shanghai before the outbreak, except that wild animal consumption was more prevalent in Wuhan than in Shanghai. During the outbreak, residents in both cities significantly changed their behaviours and reduced exposures to live animals, and such changes were much more significant in Wuhan. Most respondents supported governmental containment measures implemented, and three in four had confidence on the effectiveness of these measures. Male, the less educated and those exposing to live animals at usual were less likely to support containment measures.

In our survey, more than 20% of respondents visited wet markets at usual, which was similar in Wuhan and Shanghai, despite Shanghai being much more developed. Compared with the 30% level reported in 2013 in the same study sites,^15^ the share of respondents visiting wet markets declined by 10 percentage points, but was still high. 10% of respondents consumed products of wild animals in Wuhan, twice that reported in Shanghai. In China, there exists a long tradition of eating the meat and products of wild animals, which persists into modern times due to a false but commonly held belief that these products have therapeutic effects. ^22-23^ These beliefs and preferences for fresh food are especially popular in southern China, where Wuhan is located, but were greatly challenged and changed during the 2003 SARS outbreak in China ^9,12^. However, these practices have become popular again in recent years, due in part to the popularity of social media influencers who broadcast themselves eating wild animal products on livestream platforms. ^22^ The revival of this tradition may have set the socio-biological context for the occurrence and spread of COVID-19 in China.

We found that respondents’ risks of exposure to live animals declined significantly, reaching a very low level, and most respondents stopped wild animal consumption after this outbreak, especially in Wuhan. This indicates that the public responded quickly and adequately to the outbreak. In the survey, all respondents cared about the outbreak, and about one in five respondents in Wuhan reported starting to pay attention before Dec 31, 2019. The outbreak gradually gained intense nationwide attention,^24^ and a series of policies were implemented by the Chinese government to contain the outbreak. As of Jan 30, all provinces had initiated first-level responses to major public health emergencies.^25-26^ These actions raised public attention further and induced significant behavioural changes. A national online survey on Jan 26 showed that the vast majority of people knew of the disease and related preventive measures, 66.5% reported that they would wear masks when outside their home, and 80% cancelled travel plans during the Spring Festival due to the outbreak.^27^ The findings from both this survey and ours demonstrate behavioural change in terms of exposure to live animals or daily protection.

In addition, more strict regulations were implemented on wet markets to lower the risk of disease transmission. Wuhan instituted prohibited the sale of live poultry and wild animals starting on Jan 22. Shanghai started its seasonal closure of LPMs on Jan 25 this year. The Chinese Government further instituted nationwide bans on all forms of wild animal transactions on Jan 26.^28^ These regulations may accelerate the decrease in exposing behaviours to live animals. However, the COVID-19 epidemic occurred prior to the Chinese lunar new year, when the demand for live animals is generally high.^23^ This may produce a conflict between the public demand for live animals and regulations. Fortunately, we found that the public strongly supports the regulations on wet markets to control the outbreak. Although permanent closure of LPMs was not fully backed by the public, it was still much higher than reported during the 2013 influenza A outbreak (27.3% in Wuhan, 39.3% in Shanghai).^15,29^ The strong support for this policy offers a great opportunity to improve regulations on wet markets and change the prevalent habit of eating wild animal products in China.

We believe that the keys to quelling outbreaks of zoonotic viral diseases such as COVID-19 and preventing future outbreaks are changing the false beliefs and dangerous habits regarding eating wild animal products and strengthening regulations on wet markets. Firstly, healthy diets need to be vigorously promoted in China. Health China 2030 promotes healthy diets, but does not discuss the risks of eating wild animals. We also believe the internet and social media can be an effective channel to promote healthy eating habits and reduce the inappropriate and outdated tradition of eating wild animal products.^25,30^ Since the outbreak, spontaneous initiatives have emerged on the internet to explain the risks of and resist consuming wild animals. Secondly, live animal transactions should be more strictly regulated, and wild animal transactions should be even prohibited in the long term. While different animals should be sold separately in specific wet markets to lower risks of virus transmission, in practice, various kinds of live animals are sold together within one market, as found in Huanan Seafood Wholesale Market. Laws should be strengthened to regulate and implement the strict separation of live animals from general markets. With the currently high public support, bans on the wild animal trade should be promoted, but the government must be aware of the risk of creating an unregulated black market by instituting a ban. For live poultry and non-wild animals, better regulations may be a better option than bans without the full support of the public.^23^ The feasible measures include seasonal closure of wet markets, regular rest days, and banning leaving live animals in markets overnight.^10,15^ The above measures should be given priority in China and other countries to reduce current levels of COVID-19, as well as future threats from zoonotic diseases.

Our study has several limitations. First, there may be recall bias. We aimed to identify behavioral changes in a cross-sectional survey, and outcomes before the outbreak were measured based on participant recall. Second, our results may be affected by selection bias from telephone survey. We did attempt multiple calls to unanswered numbers to mitigate this bias, and quota sampling was introduced to get a representative sample. The anonymous phone interview addressed the concern of response bias due to fear of being seen as criticizing the government. Third, a cross-sectional design prevents us from assessing how long the effect on behavioral/intention change demonstrated in this study will last, when the outbreak is no longer fresh in people’s minds. Thus, we plan to conduct a second-round survey at the end of the outbreak to track the long-term changes.

In conclusion, exposure to live animals was common before the COVID-19 outbreak. The public responded quickly to this outbreak, and significantly reduced their exposure to live animals and stopped wild animal consumption, especially in Wuhan. They were very supportive and confident for current mitigating measures. With high public support, wet markets should be better regulated, and healthy diets, including changing the habit of wild animal consumption, should be promoted, especially for male, the less educated and those exposing to live animals at usual.

## Data Availability

Full top line results for the survey are available from H.Y. at.

## Contributors

H.Y. conceptualized the study design. Z.H., M.Q., Y.L., and J.Z. developed the survey questionnaire and collected data. Z.H., L.Liang and F.D. analyzed data. Z.H. interpreted results and wrote the manuscript. L.Lin, and H.Y. edited the manuscript.

## Ethics

The study was approved by the Institutional Review Board of the School of Public Health, Fudan University (IRB#2020-TYSQ-01-1). Verbal informed consent was obtained from all participants.

## Declaration of interests

H.Y. has received research funding from Sanofi Pasteur, GlaxoSmithKline, Yichang HEC Changjiang Pharmaceutical Company and Shanghai Roche Pharmaceutical Company. None of that research funding is related to 2019-nCov.

### Acknowledgements

We thank Mark Jit from London School of Hygiene and Tropical Medicine for his valuable comments. We are thankful to Qian Wang, Xinyu Wang, Shengyi Pan, Zihan Xu, Longfei Feng, Kaiyue Ren, Yifan Cheng, Abudukelimu Nazhakaiti, Zhiqiang Qu, Geshu Zhang, Ji Geer Guliyeerke, Qian Lv, Biao Wang, LinQing Zhou, and Cong Ma from School of Public Health, Fudan University for providing assistance with data collection.

## Data sharing statement

Full top line results for the survey are available from H.Y. at.

---

**Appendix Figure 1.**
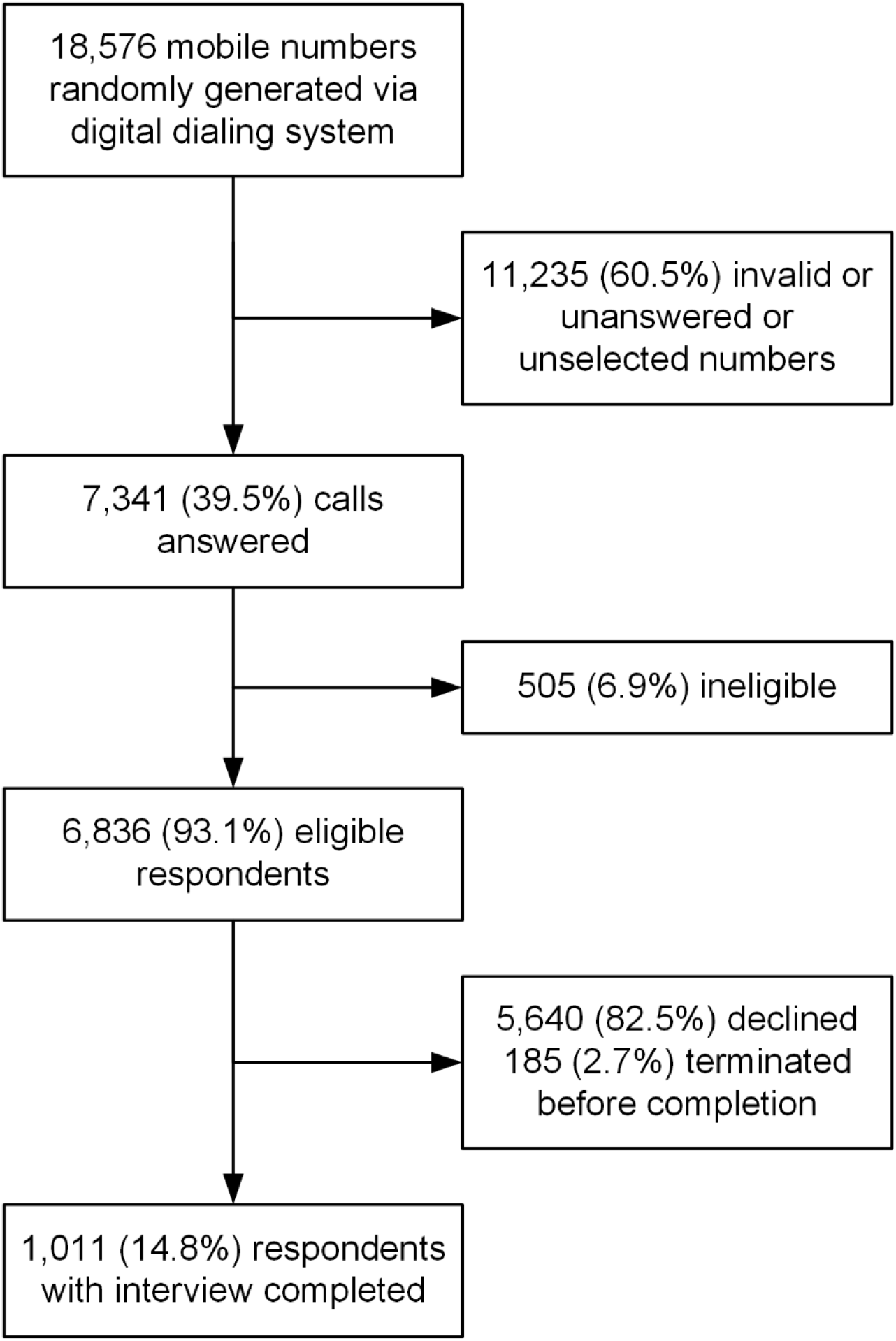
Flowchart for participant recruitment.

## Reference

1. Huang C, Wang Y, Li X, et al. Clinical features of patients infected with 2019 novel coronavirus in Wuhan, China. Lancet 2020.

2. Chen N, Zhou M, Dong X, et al. Epidemiological and clinical characteristics of 99 cases of 2019 novel coronavirus pneumonia in Wuhan, China: a descriptive study. Lancet 2020.

3. Li Q, Guan X, Wu P, et al. Early Transmission Dynamics in Wuhan, China, of Novel Coronavirus-Infected Pneumonia. N Engl J Med 2020.

4. Lu R, Zhao X, Li J, et al. Genomic characterization and epidemiology of 2019 novel coronavirus: implications for virus origins and receptor binding. Lancet 2020.

5. Zhu N, Zhang D, Wang W, et al. A novel coronavirus from patients with pneumonia in China, 2019. N Engl J Med 2020.

6. World Health Organization. Statement on the second meeting of the International Health Regulations (2005) Emergency Committee regarding the outbreak of novel coronavirus (2019-nCoV). 30 January 2020. Geneva, Switzerland.

7. National Health Commission of the People’s Republic of China. http://www.nhc.gov.cn. (Assessed on February 10th, 2020)

8. Wu JT, Leung K, Leung GM. Nowcasting and forecasting the potential domestic and international spread of the 2019-nCoV outbreak originating in Wuhan, China: a modelling study. Lancet 2020.

9. Webster RG. Wet markets--a continuing source of severe acute respiratory syndrome and influenza? Lancet 2004; 363(9404): 234–6.

10. Peiris JS, Cowling BJ, Wu JT, et al. Interventions to reduce zoonotic and pandemic risks from avian influenza in Asia. Lancet Infect Dis 2016;16(2): 252–8.

11. Guan Y, Zheng BJ, He YQ, et al. Isolation and characterization of viruses related to the SARS coronavirus from animals in southern China. Science 2003; 302(5643): 276–8.

12. Wang LF, Eaton BT. Bats, civets and the emergence of SARS. Curr Top Microbiol Immunol 2007; 315: 325–44.

13. Azhar EI, El-Kafrawy SA, Farraj SA, et al. Evidence for camel-to-human transmission of MERS coronavirus. N Engl J Med 2014; 370: 2499–505.

14. Siedner MJ, Ryan ET, Bogoch II. Gone or forgotten? The rise and fall of Zika virus. The Lancet Public Health 2018; 3(3): e109–10.

15. Wang L, Cowling BJ, Wu P, et al. Human exposure to live poultry and psychological and behavioral responses to influenza A(H7N9), China. Emerg Infect Dis 2014; 20(8):1296–305.

16. Wu P, Wang L, Cowling BJ, et al. Live Poultry Exposure and Public Response to Influenza A(H7N9) in Urban and Rural China during Two Epidemic Waves in 2013-2014. PLoS One 2015; 10(9): e0137831.

17. Liao Q, Wu P, Lam WW, et al. Public risk perception and attitudes towards live poultry markets before and after their closure due to influenza A(H7N9), Hong Kong, January-February 2014. JPublic Health (Oxf) 2016; 38(1): 34–43.

18. Kavanagh MM. Authoritarianism, outbreaks, and information politics. The Lancet Public Health 2020.

19. Rubin GJ, Amlôt R, Page L, Wessely S. Public perceptions, anxiety, and behaviour change in relation to the swine flu outbreak: cross sectional telephone survey. BMJ 2009.

20. Leung GM, Ho LM, Chan SK, Ho SY, Bacon-Shone J, Choy RY, et al. Longitudinal assessment of community psychobehavioral responses during and after the 2003 outbreak of severe acute respiratory syndrome in Hong Kong. Clin Infect Dis 2005; 40: 1713–20.

21. Leung GM, Lam TH, Ho LM, Ho SY, Chan BH, Wong IO, et al. The impact of community psychological responses on outbreak control for severe acute respiratory syndrome in Hong Kong. J Epidemiol Community Health 2003; 57: 857–63.

22. Li J, Li J, Xie X, et al. Game consumption and the 2019 novel coronavirus. Lancet Infect Dis 2020.

23. Yu H, Wu JT, Cowling BJ, et al. Effect of closure of live poultry markets on poultry-to-person transmission of avian influenza A H7N9 virus: an ecological study. Lancet 2014; 383(9916): 541–8.

24. Zhong N. Response to the prevention and control of the COVID-19. Xinhua net. http://www.xinhuanet.com/2020-01/20/c_1125487200.htm.

25. Rubin GJ, Wessely S. The psychological effects of quarantining a city. BMJ 2020; 368: m313.

26. Xiang YT, Yang Y, Li W, et al. Timely mental health care for the 2019 novel coronavirus outbreak is urgently needed. Lancet Psychiatry 2020.

27. The Paper news. Public Awareness and Prevention Behavior of COVID-19. https://www.thepaper.cn/newsDetail_forward_5635537

28. General Administration of Market Supervision, Ministry of Agriculture and Rural Affairs, National Forestry and Grass Bureau. Announcement on Prohibition of Wildlife Trading (No. 4 of 2020). Jan 26, 2020.

29. Yuan J, Liao Q, Xie CJ, et al. Attitudinal changes toward control measures in live poultry markets among the general public and live poultry traders, Guangzhou, China, January–February, 2014. Am J Infect Control 2014; 42: 1322–4.

30. Atlani-Duault L, Ward JK, Roy M, Morin C, Wilson A. Tracking online heroisation and blame in epidemics. The Lancet Public Health 2020.

